# The Role of VSL#3® in the Treatment of Fatigue and Other Symptoms in Long Covid-19 Syndrome: a Randomized, Double-blind, Placebo-controlled Pilot Study (DELong#3)

**DOI:** 10.1101/2023.06.28.23291986

**Authors:** Beatrice Marinoni, Alessandro Rimondi, Federico Bottaro, Clorinda Ciafardini, Chiara Amoroso, Martina Muià, Bruna Caridi, Daniele Noviello, Alessandra Bandera, Andrea Gori, Marco Mantero, Francesco Blasi, Roberta Ferrucci, Federica Facciotti, Maurizio Vecchi, Flavio Caprioli

## Abstract

Long COVID, also known as Post-acute COVID-19 Syndrome (PACS), is a chronic condition affecting individuals who have recovered from acute COVID-19. It is currently estimated that around 65 million people worldwide suffer from Long COVID. It is characterized by a range of symptoms, including fatigue, exertion intolerance, neurocognitive and sensory impairment, sleep disturbance, myalgia/arthralgia, and dysautonomia. Among them fatigue has emerged as a burdensome and pervasive issue, significantly impacting the quality of life and daily functioning of Long COVID patients. Alterations in the composition of the intestinal microbiota has been reported in COVID-19 patients. Dysbiosis persists even after several months of recovery from acute SARS-CoV-2 infection.

Based on this evidence, we carried out a phase 3, randomized, double-blind, placebo-controlled trial aimed at evaluating the efficacy of VSL#3®, a consortium of probiotic bacterial strains, in reducing fatigue and improving various aspects of patients’ well-being in patients with Long COVID syndrome.

**Highlights:**

- Patients suffering from Long-COVID manifest a variety of persistent symptoms impacting daily functioning;
- Fatigue emerged as a burdensome and pervasive issue, significantly impacting the quality of life;
- VSL#3® treatment significantly reduced the Chalder Fatigue Scale scores as compared to placebo
- Chalder Fatigue Scale scores remained significantly reduced in the treatment group 4 weeks post intervention.

## Introduction

Long COVID (LC) or post-COVID syndrome (PCS), is a debilitating condition affecting patients who recovered from acute COVID-19 illness ^1^. According to the World Health Organization, the global prevalence of LC is estimated to be approximately 10% of individuals after resolution of COVID-19, beyond 4 to 12 weeks after infection. As of May 2023, over 767 million people have been infected worldwide by SARS-COV-2 it is estimated that currently over 75 million individuals worldwide suffer from LC (World Health Organization. WHO coronavirus (COVID-19) Dashboard).

LC manifests with a variety of symptoms, the most prevalent being fatigue, shortness of breath, muscle pain, joint pain, headache, cough, chest pain, attention deficit (brain fog), heartburn, altered smell and taste and diarrhea ^1^. According to the largest metanalysis to date, fatigue, respiratory, and cognitive clusters occur in 51.0% (16.9–92.4), 60.4% (18.9–89.1), and 35.4% (9.4–75.1) cases of LC three months from recovery from symptomatic COVID-19 infection ^2^.

The proportion of LC cases appeared greater in individuals who were admitted to intensive care units (ICU) and general hospital wards ^3^. Hospitalized cases were estimated to experience a longer median duration of LC ^3^ and the risk to develop LC increased with age and in females ^4^.

The pathogenetic mechanisms and contributing factors to LC are likely multifaceted and potentially intersecting. Among them, sustained presence of SARS-CoV-2 in body tissues ^5^, dysregulation of the immune system ^6–8^, reactivation of pre-existing pathogens like Epstein–Barr virus (EBV) and human herpesvirus 6 (HHV-6) ^9,10^, changes in immune-metabolic pathways and microvascular clotting paired with endothelial dysfunction have been proposed.

However, the absence of clear pathogenic pathways has contributed to slow progress in the development of effective therapeutics for LC. Currently both pharmacological and non-pharmacological therapies have been assessed for specific LC symptoms ^1^. For example, L-arginine plus vitamin C supplementation demonstrated to improve walking performance, muscle strength, endothelial function, and fatigue in adults with long COVID ^11^. Two ongoing trials are examining the effects of nicotinamide riboside, a dietary supplement, (ClinicalTrials.gov Identifier: NCT04809974, NCT04604704) in reducing cognitive symptoms and fatigue by modulating the pro-inflammatory response ^12^.

SARS-CoV-2, the causative agent of COVID-19, has a strong affinity for the gastrointestinal system leading to various gastrointestinal symptoms commonly observed in individuals with COVID-19 and persisting even after the acute illness has resolved ^1^. During the acute phase, SARS-CoV-2 RNA is excreted by nearly 50% of patients in their stool samples ^13^. Additionally, a significant number of patients with LC symptoms had detectable viral antigens persisting in the gastrointestinal mucosa seven months after infection ^14^. This persistence of the virus in the intestinal tract may also have an impact on the balance of gut microbiota, potentially contributing to dysbiosis. Indeed, patients affected by COVID-19 manifest significant alterations in the composition of gut microbiota, persisting after clearance of the virus ^15, 16^.

Individuals experiencing LC symptoms also show microbial dysbiosis and reduced microbial diversity ^17, 18^. LC patients possess higher levels of *Ruminococcus gnavus* and *Bacteroides vulgatus* and lower levels of *Faecalibacterium prausnitzii* compared with non-COVID-19 pre pandemic controls, with gut dysbiosis lasting at least 14 months; low levels of butyrate-producing bacteria are strongly correlated with LC at 6 months ^17^. Interestingly, LC patients manifest increased relative abundance of opportunistic pathogens in the feces, that positively correlate with fatigue, respiratory symptoms, and neuropsychiatric symptoms, while decreased levels of other anti-inflammatory/butyrate-producing taxa negatively associate with LC ^18^.

Given the dysregulation of gut microbiota observed in COVID-19 and LC patients, the modulation of microbiota presents a promising therapeutic approach. In different respiratory infections, the use of probiotic supplementation has been shown to have a beneficial impact on the immune system ^19^ and several probiotics strains have been reported to positively influence respiratory and gastrointestinal health ^20^, alleviate influenza-like symptoms ^21^, enhance nasal innate immunity response to rhinovirus infection ^22^ and stimulate humoral immune response following oral vaccination in healthy adults ^23^.

VSL#3® (Actial Farmaceutica s.r.l.) is a multi-strain high potency probiotic mixture comprising Lactobacilli, Bifidobacteria, and Streptococcus thermophilus. In vitro studies have shown that exposure of Caco-2 cells to VSL#3®-derived supernatant positively impacts gut barrier function and repair processes ^24^.VSL#3® administration to COVID-19 patients discharged from hospital demonstrated a reduction in inflammatory markers ^25^.

Here, we performed a prospective single-center, randomized, double-blind, placebo-controlled trial aimed at investigating the potential benefit of VSL#3® supplement for the treatment of fatigue, quality of life, symptom somatization, performance status, and gastrointestinal symptoms in patients with LC.

## Methods

### Study design

This ongoing research is a phase three, single-center, randomized, double-blind, placebo-controlled trial (NCT05874089) to evaluate the efficacy of VSL#3®, a consortium of mixed probiotics at high concentration, in reducing fatigue and in ameliorating different aspects of patients’ well-being.

Patients were recruited from the LC Outpatient Clinic at Policlinico Hospital, Milan (Italy) and randomized to receive either VSL#3® or placebo for a duration of 4 weeks, 2 anonymized sachets twice a day.

The trial was performed in accordance with the of Helsinki and good clinical practice regulations after approval by the ethics committee of IRCCS Fondazione Ca’ Granda Ospedale Maggiore Policlinico di Milano (number S62043). Written informed consent was obtained from each participant before inclusion. All data were collected at Gastroenterology and Endoscopy Unit of IRCCS Fondazione Ca’ Granda Ospedale Maggiore Policlinico di Milano (Milan, Italy).

Participants were randomly assigned to receive either the treatment or the placebo by using RED-Cap (Research Electronic Data Capture) software (1:1). To consider the increased frequency of LC in females ^4^ a randomization with permutation in blocks stratified by sex was performed. Double blinding was achieved by packaging treatment and placebo in the same sealed and consecutively numbered containers with sachets similar in packaging, smell, and taste. All study participants and onsite study personnel remained masked for the treatment allocation (randomized controlled trial phase).

*Inclusion criteria*: adult patients between 18 and 65 years with a previous diagnosis of SARS-CoV-2 infection, documented by nasopharyngeal or antigenic molecular swab, Chalder Fatigue Scale (in dichotomous form) ≥ 4/11 possibly associated with signs and symptoms of LC, signs and symptoms that develop during or after SARS-CoV-2 infection, which persist for more than 4 weeks and are not reasonably explained otherwise; signs and symptoms include: fatigue, sleep disturbances, cognitive deficits (i.e. brain fogging, loss of concentration and memory, anxiety, depression), strength deficits, arthralgias and myalgias, gastroenterological alterations (reduced appetite, nausea, changes in bowel habits, abdominal pain). Not subjected to isolation or sanitary quarantine; No antibiotics treatment in the 30 days prior to the trial.

*Exclusion criteria*: use of antibiotics, probiotics, immunosuppressant/immunomodulatory drugs, opioids, antidepressants in the 30 days preceding study enrolment; cardiovascular and pulmonary disease with moderately severe organ dysfunction (NYHA>2, Borg scale ≥ 2); Decompensated endocrine and metabolic diseases (Cirrhosis with Child Pugh Turcot ≥ B, decompensated hypo/hyperthyroidism, decompensated hypoadrenalism), Diagnosis of FM, CFS/ME, and/or IBS prior to SARS-CoV-2 infection; Confirmed diagnoses of neurological pathologies, psychiatric diseases and cognitive disorders prior to SARS-CoV-2 infection; Previous confirmed diagnosis of chronic musculo-skeletal pathologies prior to SARS-CoV-2 infection; Refusal to participate in the study / refusal to process personal data; Pregnancy or breastfeeding; Addiction to alcohol or drugs in previous years; Participation in another clinical study in the previous 30 days or previous participation in this same trial; Known intolerance/hypersensitivity to the investigational drug or to the excipients of the placebo formulation.

Prohibited medications included: use of other probiotics during the trial; Use of antibiotics during the trial; use of immunosuppressant/immunomodulatory drugs, opioids, antidepressants. Substantial change of diet during the trial was considered a major protocol violation.

### Procedures

The probiotic VSL#3® (Actial Farmaceutical) is a patented consortium of 8 alive and freeze-dried Lactic acid bacteria and Bifidobacteria (Streptococcus thermophilus BT01; B. breve BB02, B. animalis subsp. lactis BL03, B. animalis subsp. lactis BI04, L. acidophilus BA05, L. plantarum BP06, L paracasei BP07, and L. helveticus BD08) at high concentration (total of 450 billion CFU/sachet) in a 4.4 gr sachet, mixtured with excipients. The placebo was packaged in 4.4 gr sachets, identical to those of the probiotic preparation. Patients with clinically relevant fatigue and diagnosis of Long-Covid Syndrome from Long Covid Outpatient Clinics in Policlinico di Milano Hospital were referred for a preliminary assessment of the inclusion and exclusion criteria. After screening and assessment by a single physician (BM) at screening visit (visit1; t-2), a run-in period of two weeks took place. Study procedures were done at baseline (visit 2; t0), after 4 weeks of treatment with probiotics or placebo (visit 3; t4), and after 4 additional weeks of follow-up after the end of treatment (visit 4; t8). Eligible patients were randomized to placebo or probiotics at the dosage of 2 sachets/die for 28 days. Treatment compliance was established by counting sachets and defined as good if 80% or more were used after each treatment phase.

### Study endpoints

The primary endpoint of the study was a reduction in fatigue as measured by the Chalder Fatigue Scale score after 4 weeks of VSL#3® treatment. Secondary outcomes included variations in scores of the Hospital Anxiety and Depression Scale (HADS), Short Form Health Survey (SF-36), Structured Assessment of Gastrointestinal Symptoms Scale (SAGIS), somatization dimension of Symptoms Check List-90 (SCL-90), Karnofsky Performance Status (KPS), and Visual Analogue Scale (VAS) after 4 weeks of treatment.

### Structured questionnaires

The structured questionnaires were completed at V2(t0), V3 (t4) and V4 (t8) and included: Chalder Fatigue Scale (CFS); Hospital Anxiety and depression symptoms (HADS-A and D); Short Form-36 (SF36); Symptoms Check list (SCL-12); Karnofksy Performance Status; Visual Analogue Scale (VAS); Structured Assessment of Gastrointestinal Symptoms (SAGIS).

#### Chalder Fatigue Scale (CFS)

CFS consists in 11 items. Fatigue is defined clinically relevant for scores higher than 4 out of 11. In the Likert scoring system, a total score of 11 out of 33 indicates clinically relevant fatigue ^26^

#### Hospital Anxiety and Depression Scale (HADS)

HADS is composed of 14 items, 7 of which are related to anxiety (-A) and 7 to depression (-D). Each item is scored from 0 to 3. The scores of each domain are reported as the arithmetic mean of the scores of the symptoms composing the domain in each subject. The final score ranges from 0 to 21 with a cutoff value ≥11 suggestive of the condition

#### Short Form-36 (SF)-36

(SF)-36 consists of 36 items that measure eight different domains: physical functioning, role limitations due to physical problems, bodily pain, general health perceptions, vitality, social functioning, role limitations due to emotional problems, and mental health. Scoring the (SF)-36 involves transforming raw item responses into standardized scores ranging from 0 to 100 for each domain. Higher scores indicate better health-related quality of life in the respective domain. ^28^.

#### Symptom Checklist (SCL)-12

(SCL)-12 includes 12 questions (headache, faintness or dizziness, nausea or upset stomach, pains in the heart or chest, pains in the lower back, soreness of your muscles, trouble getting your breath, hot or cold spells, numbness or tingling in parts of your body, a lump in your throat, feeling weak in parts of your body, heavy feelings in your arms or legs) scored on how much that problem bothered or distressed during the past week (from 0: not at all to 4: extremely). The individual raw scores were converted to standard area t-scores based on a non-psychiatric patient normative sample ^29^.

#### Karnofsky Performance Status (KPS)

KPS consists in a 11-point scale correlating to percentage values ranging from 100% (no evidence of disease, no symptoms) to 0% (death). The KPS considers various factors, including physical abilities, self-care, mobility, and the need for assistance ^30^.

#### Visual Analogue Scale (VAS)

The Visual-Analogue Scale (VAS) is a subjective rating scale allowing patients to express their subjective assessment of their overall well-being. The VAS consists of a horizontal line that ranges from one extreme to another, with descriptive anchors at each end. Patients are asked to place a mark on the line to indicate their current state of health, with one end representing poor health and the other end representing excellent health. The distance between the left end of the line and the patient’s mark is then measured to obtain a numerical rating that quantifies the patient’s self-perceived general state of health.

#### Structured Assessment of Gastrointestinal Symptoms (SAGIS)

SAGIS questionnaire includes 22 gastrointestinal symptoms scored on a five-point Likert scale as 0: no problem, 1: mild (a symptom can be ignored when you do not think about it) 2: moderate (it cannot be ignored, but does not influence daily activities), 3: severe (influencing your concentration on daily activities), and 4: very severe (it markedly influences your daily activities and/ or requires rest). Symptoms were grouped in five symptoms domains: 1-Abdominal pain/discomfort including: post-prandial pain, epigastric pain, bloating, fullness, early satiety, retrosternal discomfort, and abdominal cramps. 2-Diarrhea/incontinence including: diarrhea, loose stools, urgency to defecate, pain/ discomfort prior to defecation, excessive gas flatulence, and incontinence. 3-Gastroesophageal reflux disease/regurgitation including: dysphagia, excessive belching, and acid eructation. 4-Nausea/vomiting including: sickness, nausea, vomiting, and loss of appetite. 5-Constipation including: constipation and difficult defecation. The scores of each symptom domain are reported as the arithmetic mean of the scores for the symptoms of the given domain. The SAGIS questionnaire also asked subjects to describe in their own words their first and second most important health concern/problem and the presence/absence of 6 selected non-gastrointestinal symptoms including: headache, back pain, sleep disturbances, chronic fatigue, and self-reported depression and anxiety

### Statistical Analysis

All statistical analyses were performed using RStudio (R Core Team (2020). R: A language and environment for statistical computing. R Foundation for Statistical Computing, Vienna, Austria. URL https://www.R-project.org/.) and GraphPad Prism 9.

For continuous variables, comparison between groups were performed using the Student’s T Test and the Wilcoxon Mann Whitney Test, in the case of parametric and non-parametric tests respectively. In the case of in the case of categorical variables, the Chi-Square Test or Fisher’s Exact Test were used in the case of parametric and non-parametric tests, respectively. P-value p<0.05 was statistically significant.

## Results

### Demographic characteristics of the population

From November 3, 2022, through May 9, 2023, a cohort of 279 patients who suffered from COVID-19 infection and receiving a follow-up for persisting symptoms were evaluated for inclusion in the trial. A total of 39 individuals, presenting with clinically significant fatigue, were selected for participation. The significant level of fatigue was determined by a Chalder Fatigue Scale (CFS) score exceeding 4 out of 11. Following enrollment, one participant was dismissed due to low adherence to the study protocol. Participants were randomly divided into two groups: 19 were administered the probiotic VSL#3®, while the other 19 were given a placebo for a duration of 4 weeks, 2 anonymized sachets twice a day (**Figure 1**).

**Figure 1.**
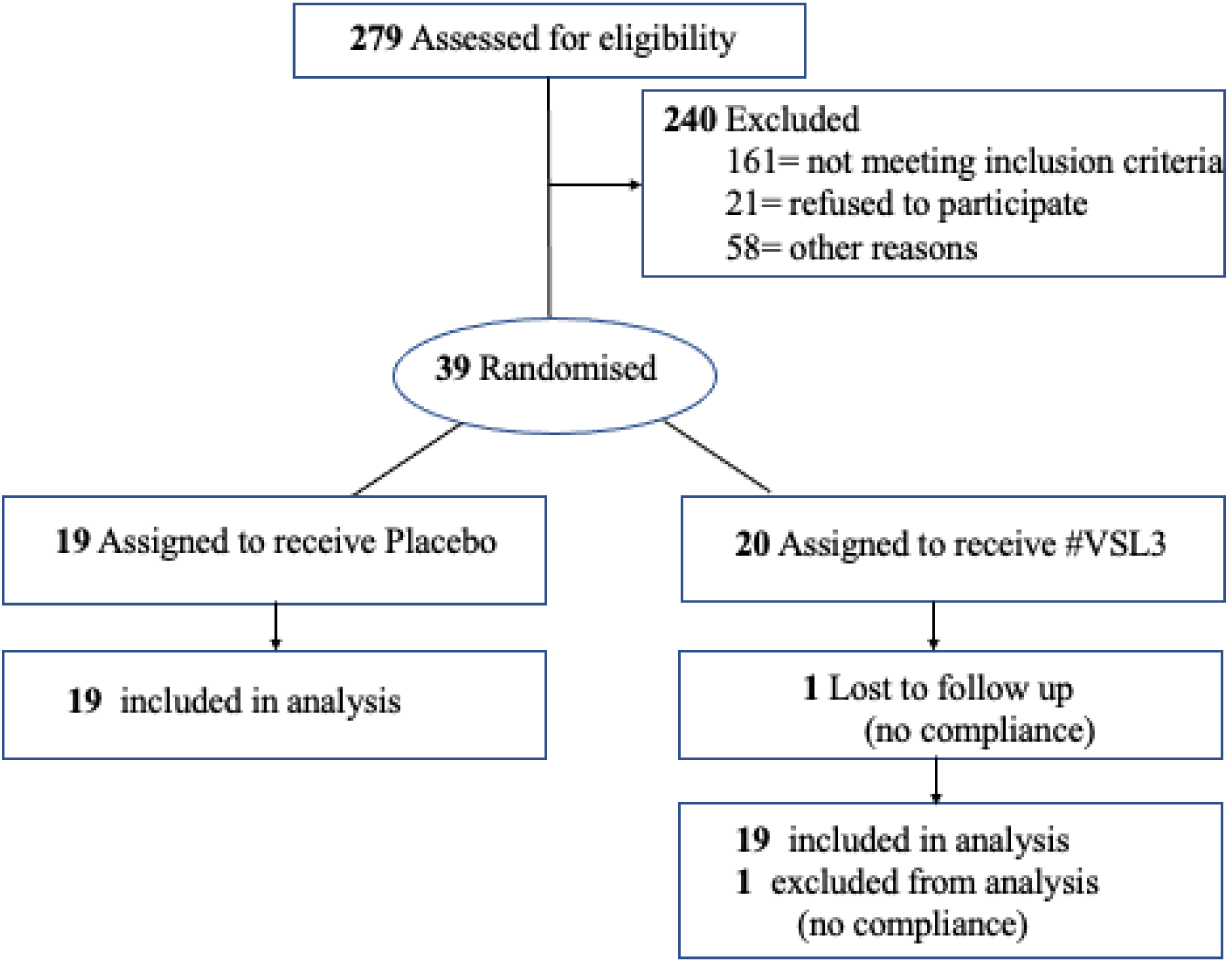
Study flow chart.

As detailed in **Table 1**, the sample consisted of 20 males (47.4%), with an average age of 54.2 years (±7.94) and an average Body Mass Index (BMI) of 26.29 (±5.39). Of the participants, 23 (60.5%) had experienced acute COVID-19 associated with interstitial pneumonia, and 22 (57.9%) had required hospitalization, with a mean length of stay totaling 13.1 days (±16.75). Among the hospitalized patients, 11 (28.9%) received Continuous Positive Airway Pressure (C-PAP) therapy, while 7 (18.4%) required admission to the Intensive Care Unit (ICU) and received tracheal intubation (ETI).

**Table 1.** Baseline clinical characteristics of the enrolled subjects.

| **Clinical parameter** | **Total**<br>**(n=38)** | **Placebo**<br>**(n=19)** | **VSL#3®**<br>**(n=19)** | **p-value** |
| --- | --- | --- | --- | --- |
| Male/Female, n (%) | 20/18 (47.4) | 11/8 (42.1) | 9/10 (52.6) | 0.52 |
| Age at enrolment, mean ± SD (years) | 54.16 (±7.9) | 54.47 (±9.4) | 53.84 (±6.5) | 0.81 |
| Body mass index BMI, mean (± SD) | 26.29 (±5.4) | 27.05 (±6.4) | 25.53 (±4.1) | 0.39 |
| Current smoker, n (%) | 6 (15.8%) | 2 (10.52) | 4 (21.05) | 0.37 |
| Current drinker, n (%) | 13 (34.2%) | 9 (47.36) | 4 (21.05) | 0.09 |
| Hospitalization, n (%) | 22 (57.9) | 11 (57.9) | 11 (57.9) | 1 |
| Hospitalization, days (± SD) | 13.1 (±16.8) | 16.55 (±29.4) | 20 (±38.4) | 0.48 |
| ICU, n (%) | 7 (18.4) | 4 (21.1) | 3 (15.8) | 0.68 |
| ETI, n (%) | 7 (18.4) | 4 (21.1) | 3 (15.8) | 0.68 |
| C-PAP, n (%) | 11 (28.9) | 6 (31.6) | 5(26.3) | 0.72 |
| Interstitial pneumonia, n (%) | 23 (60.5) | 12 (31.6) | 11 (28.9) | 0.75 |
| Abx_acute infection, n (%) | 22 (57.9) | 12 (63.2) | 10 (52.6) | 0.51 |
| Antivirals_acute infection | 12 (31.6) | 4 (21.1) | 8 (42.1) | 0.16 |
| Acute_GI symptoms | 15 (39.5) | 7 (36.8) | 8 (42.1) | 0.74 |
| CFS Score at Baseline, mean (± SD) | 23 (±4.7) | 22.4 (±4.5) | 23.0 (±4.6) | 0.31 |
| LC Length, months (± SD) | 27.0 (±10.7) | 26.5 (±10.7) | 27.7 (±10.2) | 0.21 |
| LC Symptoms |  |  |  |  |
| Brain Fogging (%) | 37 (97.4) | 19 (100) | 18 (94.7) | 0.31 |
| Gastrointestinal (%) | 28 (73.7) | 17 (89.4) | 11 (57.8) | 0.03 |
| Concentration (%) | 35 (92.1) | 16 (84.2) | 19 (100) | 0.07 |
| Memory Loss (%) | 26 (68.4) | 11 (57.9) | 15 (78.9) | 0.16 |
| Dyspnea (%) | 19 (50) | 8 (42.2) | 11 (57.9) | 0.33 |
| Chest pain (%) | 1 (2.6) | 0 | 1 (5.3) | 0.31 |
| Cough (%) | 4 (10.5) | 2 (10.5) | 2 (10.5) | 1 |
| Anosmia (%) | 0 | 0 | 0 | 1 |
| Dysgeusia (%) | 0 | 0 | 0 | 1 |
| Headache (%) | 7 (18.4) | 3 (15.8) | 4 (21.1) | 0.68 |
| Rhinitis (%) | 2 (5.3) | 1 (5.3) | 1 (5.3) | 1 |
| Alopecia (%) | 5 (13.2) | 3 (15.8) | 2 (10.5) | 0.63 |
| Poor Appetite (%) | 6 (15.8) | 2 (10.5) | 4 (21.1) | 0.37 |
| Dizziness (%) | 16 (42.1) | 8 (42.1) | 8 (42.16) | 1 |
| Myalgias (%) | 26 (68.4) | 12 (63.2) | 14 (73.7) | 0.48 |
| Joint pain (%) | 26 (68.4) | 11 (57.9) | 15 (78.9) | 0.16 |
| Sweating (%) | 1 (2.6) | 0 | 1 (5.3) | 0.31 |

SD, standard deviation; ICU, Intensive Care Unit; ETI Endotracheal Intubation; C-PAP Continuous Positive Airway Pressure; Abx, antibiotics; GI, gastrointestinal; CFS, Chalder Fatigue Scale; LC, Long Covid

**Table 2:**
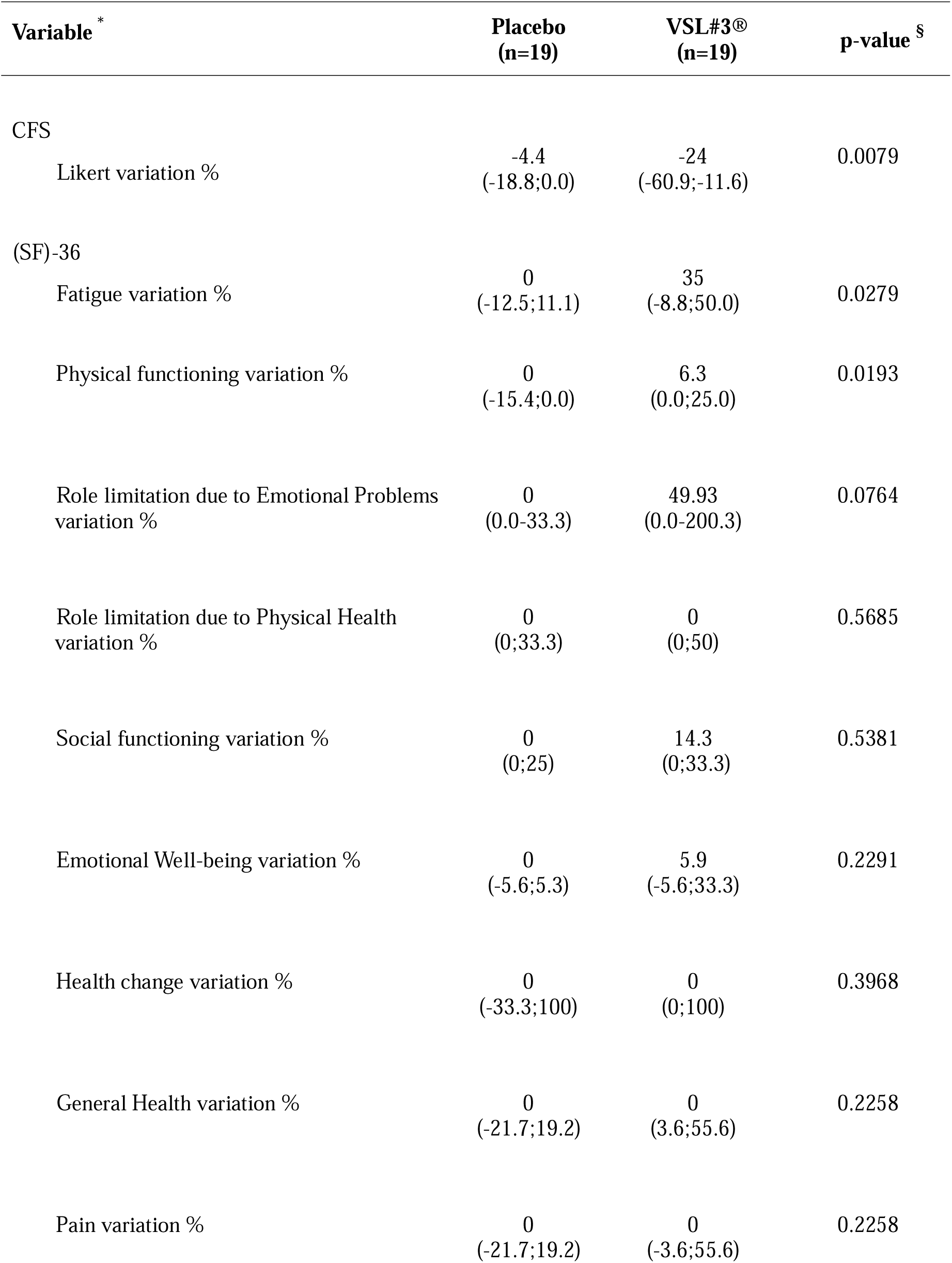
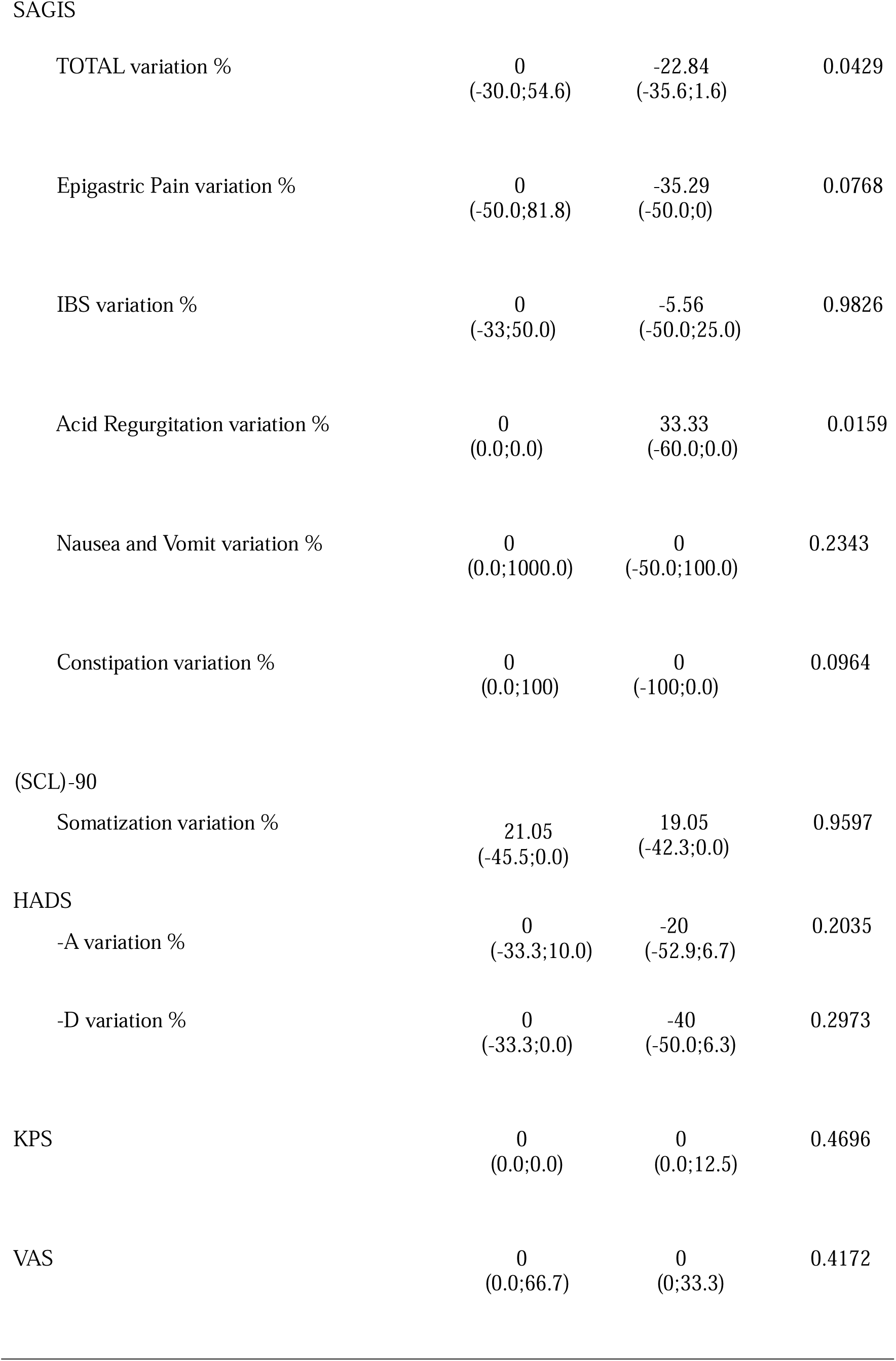
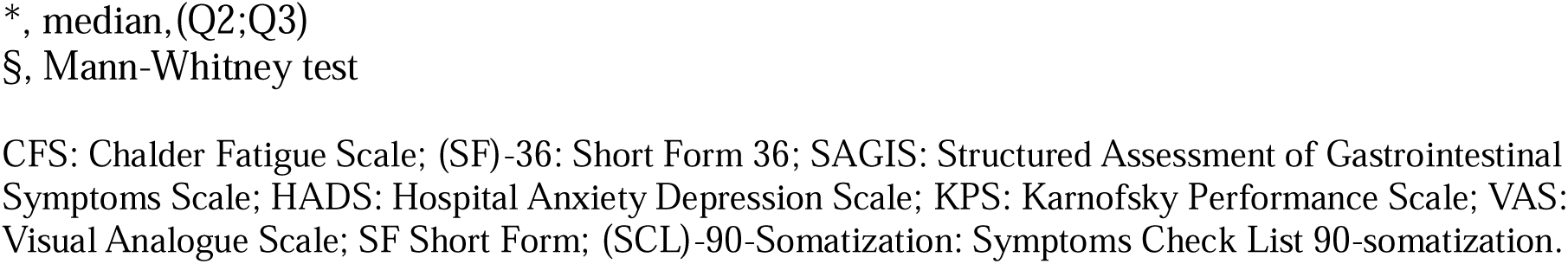
Statistical Analysis.

All enrolled patients developed persistent symptoms characteristic of Long-COVID following recovery from the acute phase of COVID-19. The spectrum of symptoms among the participants included: cognitive impairment or ’brain fog’ (97.4%), diminished concentration (92.1%), gastrointestinal complications (73.7%), memory loss (68.4%), musculoskeletal pain (68.4%), dizziness (42.1%), shortness of breath or dyspnea (50%), poor appetite (unspecified %), hair loss or alopecia (13.2%), chronic cough (10.5%), chronic rhinitis (5.3%), and chest discomfort (2.6%).

When comparing characteristics of the groups, the age, and the ratio of male to female did not significantly differ between the VSL#3® and the placebo group, thus confirming that randomization was successful. Body mass index, rate of hospitalization, severity of acute COVID-19, accompanying symptoms of LC and other health-related characteristics such as cigarette use, alcohol use, and self-evaluated health status, were not different between the placebo and VSL#3® group. No clinically relevant adverse events were reported during the intervention.

### VSL#3® Treatment significantly reduced Fatigue in Long COVID patients

The primary study endpoint was to evaluate the efficacy of VSL#3^®^ in comparison to a placebo in reducing LC-associated fatigue as assessed by Chalder Fatigue Scale, after a treatment period of four weeks. A highly significant difference between the group receiving VSL#3^®^ compared to placebo was reported in the reduction of Chalder Fatigue Score, measured using the Likert scale, from the start of treatment (t0) to its conclusion (t4)(**Figure 2A**). Of note, the significant reduction in Chalder Fatigue Score persisted 4 weeks post treatment, at the end of the study (timepoint t8, **Figure 2A**). Importantly, fatigue amelioration at the end of the trial was observed only in patients treated with the VSL#3^®^ and not in those treated with placebo (**Figure 2B**). These results indicate that the trial successfully met its primary endpoint and treatment beneficial effects persisted at least 4 weeks following treatment discontinuation.

**Figure 2:**
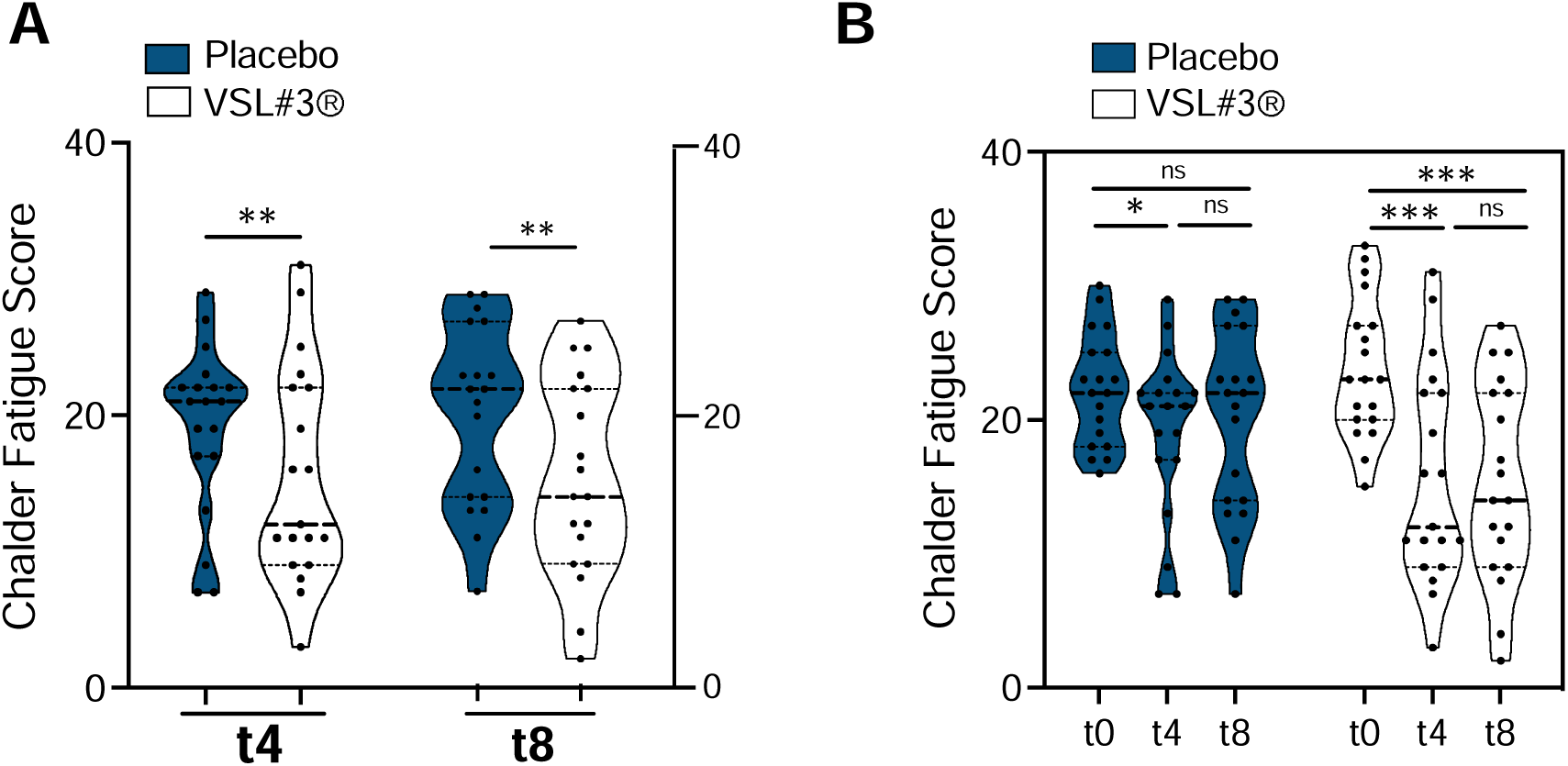
*Fatigue variation measured by Chalder fatigue Score*. Percentage variation of the different domains of the Chalder Fatigue score measured at enrollment (t0), at the end of supplementation (t4) and 4 weeks after the end of the treatment (t8) in placebo-(blue graphs) or VSL#3®-treated subjects (pink graphs). Score has **A**, Fatigue score variation between placebo and VSL#3®-treated at t4 and t8; **B**, longitudinal variation of the fatigue score in placebo and VSL#3®-treated subjects at t0, t4 and t8. Statistical analysis has been performed by Mann Whitney test, two tailed. P<0.05 was considered statistically significant. ns= not significant.

### VSL#3® Treatment ameliorates Fatigue and Physical Functioning QoL domains of the SF-36

Quality of life was assessed by the Short Form-36, which evaluates physical functioning, role limitations due to physical problems, role limitations due to emotional problems, fatigue, emotional well-being, social functioning bodily pain and general health perceptions ^28^. Any of the eight domains of the (SF)-36 were assessed before and after treatment with VSL#3® or placebo (**Figure 3**)

**Figure 3:**
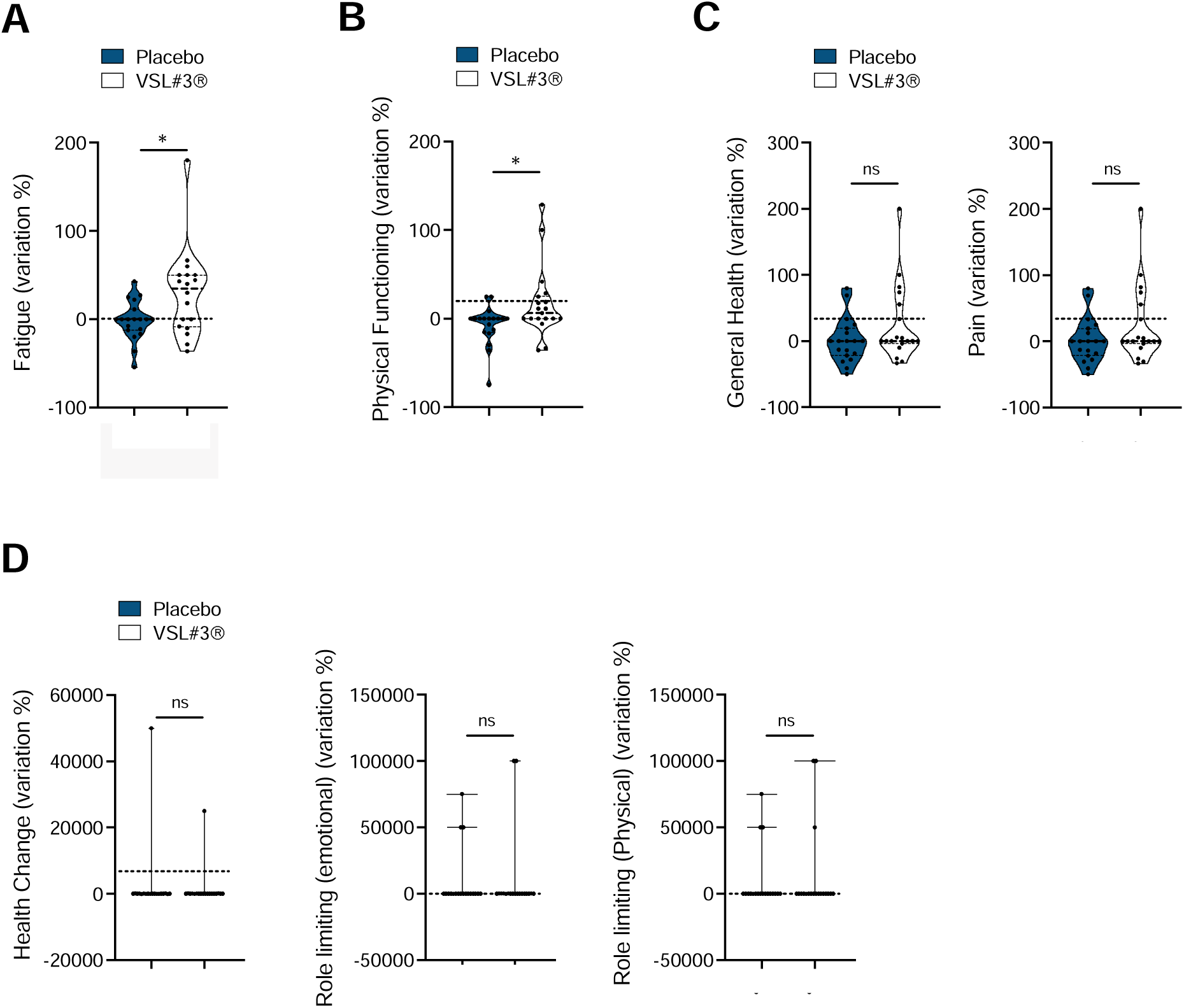
Health-related quality of life measured by SF-36 score variation. Percentage variation of the different domains of the SF-36 scoring system measured at enrollment (t0) and at the end of supplementation (t4) in placebo-(blue graphs) or VSL#3®-treated subjects (pink graphs). **A**, fatigue domain p=0.0279; **B**, physical functioning, p=0.0193; **C,** General health (left panel) and pain (right panel) **D**, health change (left panel), role limitations due to emotional problems (middle panel), role limitations due to physical problems (right panel). Statistical analysis has been performed by Mann Whitney test, two tailed. P<0.05 was considered statistically significant. ns= not significant.

The data confirm a statistically significant decrease in the Fatigue domain from the start of treatment (t0) to its conclusion (t4) in the group receiving VSL#3® compared to those receiving the placebo (**Figure 3A**). Interestingly, also the Physical functioning domain showed a significant amelioration after treatment with VSL#3® (**Figure 3B**). The remaining domains of (SF)-36 were not significantly different in placebo group and in the VSL#3® treated groups after 4 weeks of treatment (**Figure 3C,D**).

These data confirm the positive effect of VSL#3® on LC fatigue-associated symptoms.

### VSL#3® Treatment ameliorates gastrointestinal symptoms

Individuals suffering from Long COVID exhibit increased risk of incident gastrointestinal disorders spanning several disease categories including motility disorders, acid related disorders (dyspepsia, gastroesophageal reflux disease, peptic ulcer disease), functional intestinal disorders, acute pancreatitis, and hepatic and biliary disease ^32^.

Structured Assessment of Gastrointestinal Symptoms Scale (SAGIS) was assessed before and after treatment with VSL#3® or placebo (**Figure 4**). The amelioration of Gastrointestinal Symptoms from the start of treatment (t0) to its conclusion (t4), was notably more pronounced in the group receiving VSL#3® compared to those receiving the placebo. This difference was statistically significant (**Figure 4A**). Specific aspects, including acid regurgitation (**Figure 4B**), nausea and vomiting, constipation, epigastric pain and IBS symptoms (**Figure 4C**) did not vary between treated and placebo arms.

**Figure 4:**
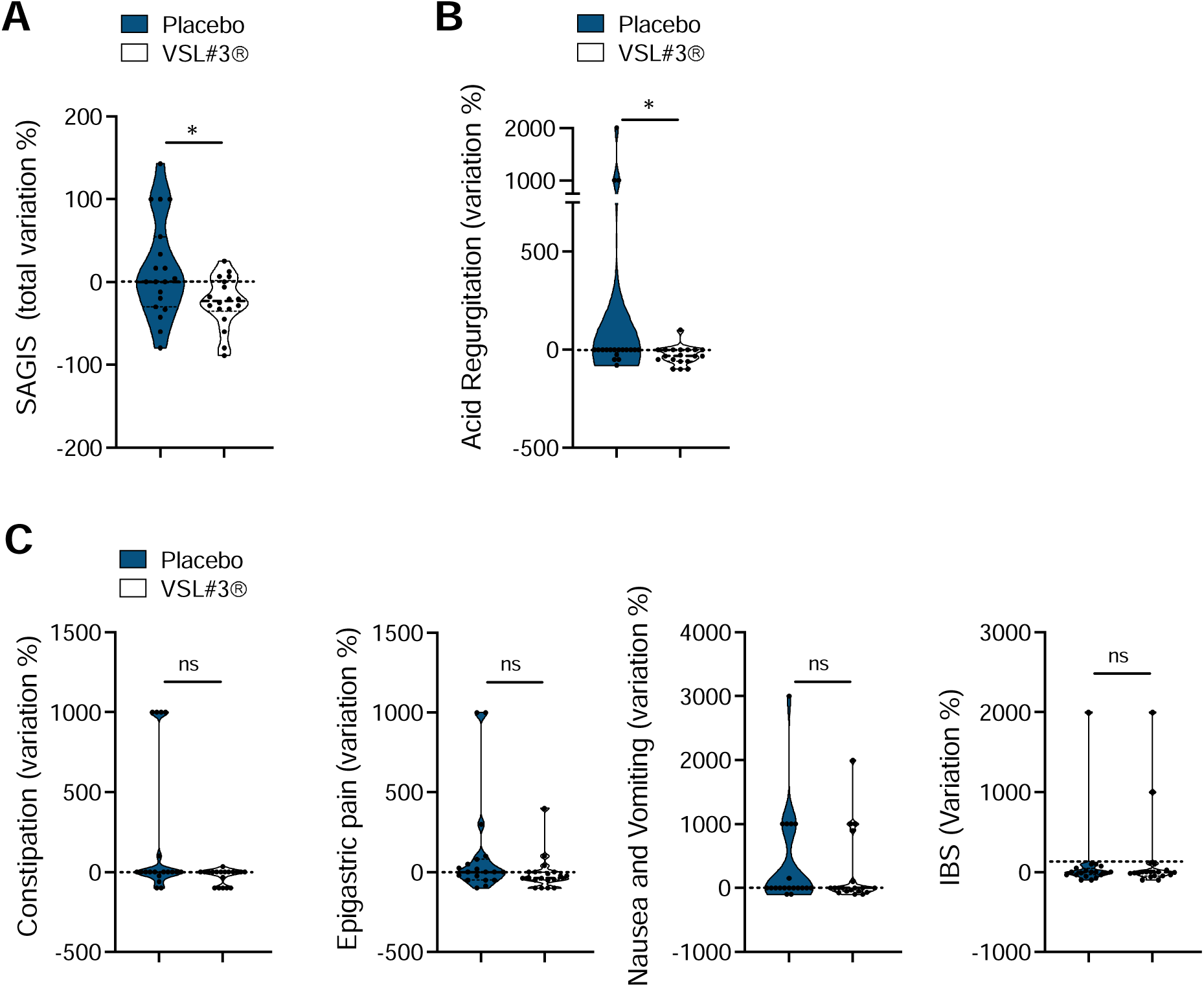
*Gastrointestinal symptoms measured by SAGIS score variation*. Percentage variation of the different domains of the SAGIS scoring system measured at enrollment (t0) and at the end of supplementation (t4) in placebo-(blue graphs) or VSL#3®-treated subjects (pink graphs). **A**, total SAGIS score p=0.04299; **B**, Acid Regurgitation 0.0159; **C,** Constipation, Epigastric pain, Nausea and vomiting, IBS symptoms. Statistical analysis has been performed by Mann Whitney test, two tailed. P<0.05 was considered statistically significant. ns= not significant

These data indicate that VSL#3® generally ameliorates gastrointestinal symptoms in Long-COVID patients.

### VSL#3® Treatment does not ameliorate Anxiety, Depression, performance and somatization of symptoms

Incidence rates and relative risks of neurological and psychiatric diagnoses are increased in patients within 6 months following a COVID-19 diagnosis ^33^. Here, the possible variation in the levels of Hospital Anxiety and Depression Scale (HADS) Scores after VSL#3® treatment were assessed. The data indicate that neither HADS-A nor HADS-D significantly differ from the start of treatment (t0) to its conclusion (t4) in the group receiving VSL#3® compared to those receiving the placebo (**Figure 5A**, left and right panels).

**Figure 5:**
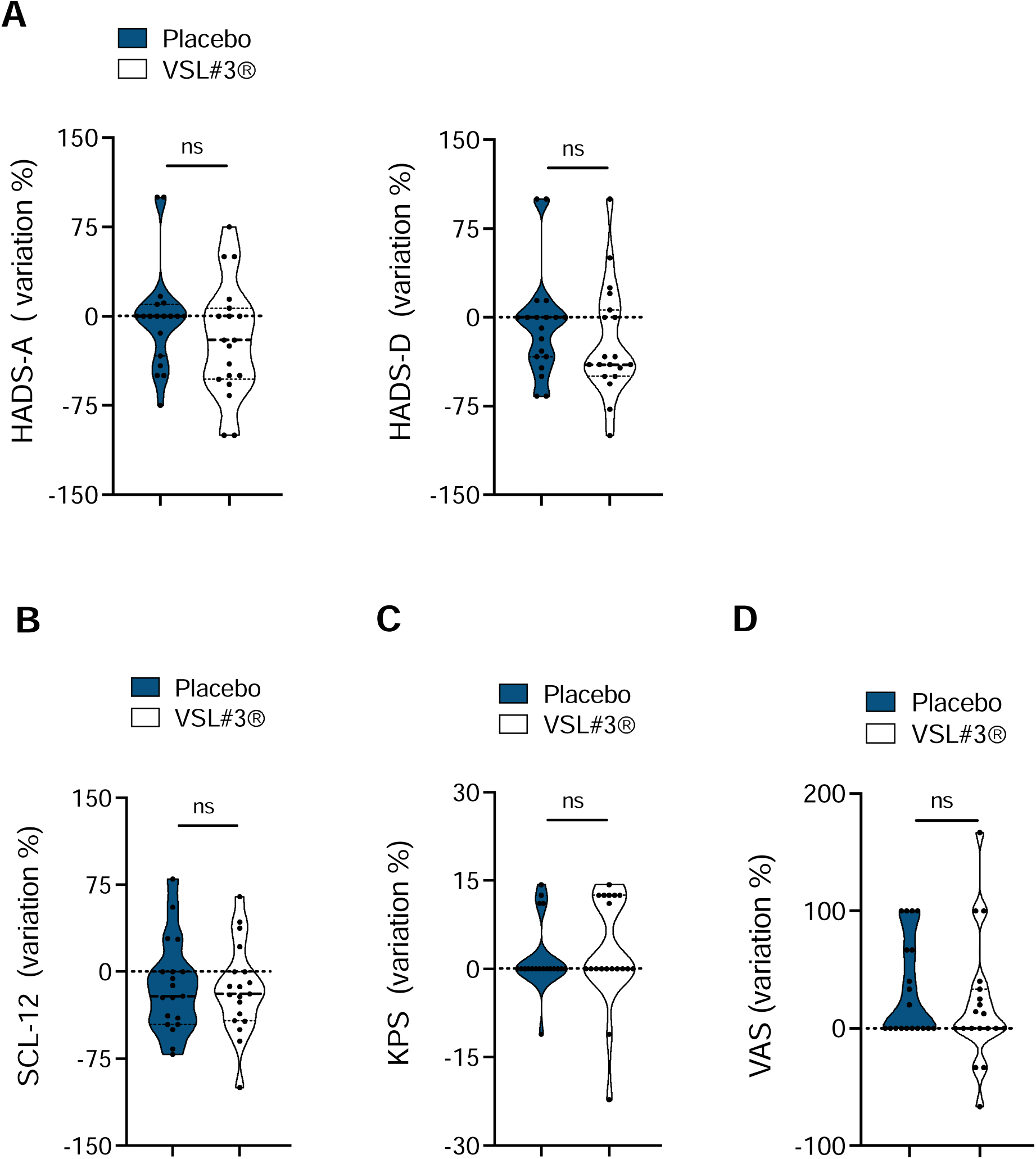
*Mental health and physical functioning variation*. Percentage variation of the different domains of the HADS, SCL-12, KPS and VAS scoring system measured at enrollment (t0) and at the end of supplementation (t4) in placebo-(blue graphs) or VSL#3®-treated subjects (pink graphs). **A**, HADS-A (left panel) and HADS-D (right panel); **B**, SCL-12; **C** KPS, **D,** VAS. Statistical analysis has been performed by Mann Whitney test, two tailed. ns= not significant

Patients suffering from LC report a variety of symptoms impairing daily life, including headache, faintness or dizziness, nausea, pain, sore muscles, shortness of breath, hot or cold spells, numbness or tingling in body parts, arms, or legs weakness (1). The Symptom Check-List 12, (SCL)-12 collects the reports of these and other symptoms, while the Karnofsky Performance Status Scale and the Visual Analogue Scale collect information concerning performance and patient’s overall functional ability and daily activity.

The analyses at t4 indicate that none of these values differ in the group receiving VSL#3® compared to those receiving the placebo (**Figures 5B-D**).

The data indicate that supplementation of VSL#3® does not significantly ameliorate symptoms reported in the HADS-A and -D, SCL-12, KPS and VAS scales.

## Discussion

In this study we conducted a prospective, randomized, double-blind, placebo-controlled trial to investigate the potential benefits of VSL#3® supplementation in patients with Long COVID, with the goal of reducing symptoms of fatigue. Our results, although preliminary, indicate that VSL#3® supplementation is beneficial in reducing fatigue symptoms and improving gastrointestinal symptoms in patients with Long COVID. Remarkably, the primary outcome measure, the Chalder Fatigue Scale (CFS), showed a statistically significant difference between the placebo and treated groups at the end of the treatment period (t4), which was maintained also for additional 4 weeks post treatment (t8).

Gastrointestinal symptoms were additionally ameliorated upon VSL#3® supplementation. These results support the notion that targeting the gut microbiota with VSL#3® may have a positive impact on fatigue, aspects of quality of life, and gastrointestinal symptoms in patients with Long COVID. These results are also consistent with previous studies indicating a link between gut dysbiosis and fatigue ^30, 31^.

Chronic fatigue syndrome (CFS) is a complex and debilitating disorder characterized by persistent fatigue that is not relieved even by rest. It is often accompanied by cognitive impairment, sleep disturbances, and musculoskeletal pain. While the exact etiology of CFS remains unclear, there is evidence that viral infections, including SARS-CoV-2, may trigger or exacerbate fatigue symptoms^34^. Reactivated herpes virus infections, primarily EBV and HHV-6, have been implicated in severe COVID-19 and the formation of the Long COVID syndrome ^9^. The same viruses have also been identified in Myalgic Encephalo-mylitis / Chronic Fatigue Syndrome (ME/CFS) cases ^35^. These reactivated viruses can cause mitochondrial fragmentation and significantly disrupt energy metabolism ^36^. A recent study in preprint stage suggests a correlation between EBV reactivation and fatigue and neurocognitive dysfunction in LC patients ^10^.

The gut-brain axis, involving bidirectional communication between the gut microbiota and the central nervous system, plays a critical role in regulating various physiological processes, including mood, cognition, and immune function ^34^. which is characterized by an imbalance in the composition of the gut microbiota and has been linked to various health conditions, including fatigue and neuroinflammation.

Current research indicates that the composition of gut microbiota is significantly altered in patients affected by Long COVID ^15^. Microbial dysbiosis, however, and altered gut permeability have been reported in association with Long COVID, suggesting a possible involvement of the gut microbiota in the persistence of symptoms ^16, 37^. In a study conducted by Yeoh et al., analysis of microbiota in 100 hospitalized patients with COVID-19 revealed a notable decrease in Bifidobacteria and Lactobacilli, which stratified with disease severity and correlated with elevated concentrations of inflammatory cytokines and blood markers (i.e. C reactive protein). Interestingly, the dysregulated gut microbiota composition in patients with COVID-19 persisted after clearance of the virus ^16^. Specific taxa including *Ruminococcus gnavus* and *Bacteroides vulgatus* resulted increased while *Faecalibacterium prausnitzii* levels are decreased in people experiencing LC symptoms, with dysbiosis lasting at least 14 months ^37^. Low levels of butyrate-producing bacteria are strongly correlated with LC at 6 months. Importantly, patients who had COVID-19 but did not develop LC did not display the same pattern of dysbiosis and showed recovered microbiome profile at 6 months comparable to that of non-COVID-19 controls. Conversely, in the LC subgroup, increased relative abundance of opportunistic pathogens in the feces positively correlated with fatigue, respiratory symptoms, and neuropsychiatric symptoms, while decreased levels of other anti-inflammatory/butyrate-producing taxa were negatively associated with LC at the six-month mark ^15^. In another recent study Zhuo et al. reported reduced diversity in the microbiota of a cohort consisting of 15 patients, followed up for three months, all of whom had at least one persistent COVID-19 symptom ^13^.

Given the dysregulation of gut microbiota observed in COVID-19 and LC, the modulation of microbiota presents a promising therapeutic approach that could benefit patients through different mechanisms such as the production of short-chain fatty acids, including butyrate, which possesses anti-inflammatory effects, the modulation of the gut-brain axis and the restoration of gut barrier function by increasing the expression of tight junction proteins.

Probiotics, such as VSL#3®, offer a promising approach to manage LC symptoms by modulating the gut microbiota composition and function. By targeting dysbiosis and promoting a favorable microbial profile, probiotics have the potential to exert anti-inflammatory effects and modulate the gut-brain axis, thereby alleviating fatigue symptoms ^38^ VSL#3® is a patented consortium of live and freeze-dried lactic acid bacteria and bifidobacterial ^24, 25^

Currently the modulation of microbiota is being investigated as a potential adjunctive therapy for acute COVID-19. A published study explored the impact of probiotics on disease progression in 28 patients with acute COVID-19. These patients received antibiotics, tocilizumab, hydroxychloroquine alone or in combination, along with the administration of a multi-strain probiotic formulation (SivoMixx™) containing Lactobacillus acidophilus, L. helveticus, L. paracasei, L. plantarum, L. brevis, Bifidobacterium lactis, and Streptococcus thermophilus (2,400 billion bacteria/day). Within three days of probiotic supplementation, all patients experienced remission of diarrhea and resolution of other symptoms, in comparison to 42 healthy controls. After seven days, the probiotic group exhibited a significant reduction in the estimated risk of respiratory failure, as well as hospitalizations in intensive care units and mortality rates Multiple clinical trials are currently registered on ClinicalTrials.gov and underway to evaluate the impact of probiotics and gut microbiota modulators on COVID-19. Some aim at evaluating the effect of daily oral administration of a probiotic mixture on symptom improvement, duration of hospitalization, and viral clearance in COVID-19 patients (NCT04390477). Intranasal administration twice-daily of L. lactis (NCT04458519), or oral daily supplementation with of L. plantarum and P. acidilactici CECT 7483 in mild COVID-19 patients. (NCT04517422) are being tested. Preventive oral administration of probiotics on healthcare professionals exposed to COVID- 19 (NCT04366180) (NCT04399252) are also being evaluated. A study of synbiotic oral treatment with 8 probiotic strains and prebiotics (inulin and FOS) aims at reducing the duration of diarrhea, improve stool consistency, mitigate intestinal inflammation and dysbiosis, and alleviate gastrointestinal symptoms associated with COVID (NCT04420676).

However, controlled trials assessing microbiota modulators for LC are scarce. An ongoing study single-blind randomized placebo-controlled trial conducted by Siew et al. in Hong Kong is assessing the role of a symbionts compared to vitamin C in modulating gut microbiome and immunological response in long COVID patients (ClinicalTrials.gov Identifier: NCT04884776). Another clinical trial is currently assessing the effectiveness of a symbiotic (Omni-Biotic Pro Vi 5) supplement to normalize the composition of the gut microbiome and reduce inflammation in long COVID (ClinicalTrials.gov Identifier: NCT04813718).

Therefore, our study is the first trial aiming to specifically target fatigue as one of the most disabling symptoms of Long COVID by utilizing a probiotic consortium with high concentrations of beneficial bacteria. The objective is to reduce Long COVID symptoms, particularly fatigue, by restoring a healthy gut microbial balance (eubiosis).

Taken together, results from the present study demonstrate a significant reduction in fatigue and physical functioning upon oral administration of VSL#3® for four weeks as compared to the placebo group. This is consistent with the proposed role of probiotics in modulating the gut microbiota and their potential impact on gut-brain axis. However, further work is needed to confirm the results of this pilot study, as well as to exploit mechanisms involved in these observations. Longitudinal studies with extended follow-up periods are also warranted to assess the sustainability of the observed effects and evaluate the long-term outcomes.

## Data Availability

All data produced in the present study are available upon reasonable request to the authors

## Acknowledgments

The Authors thank Dario Consonni for help with statistical analysis, Andrea Costantino with patient recruitment, Sara Comparetti for administrative assistance.

## Role

Concept and design: FC, BM, AR, FF, DN; Acquisition, analysis, or interpretation of data: All authors, Drafting of the manuscript: BM, FC, FF; Patients recruitment, AB, AG, MM, FB, LT, MV Critical revision of the manuscript for important intellectual content: All authors. Administrative, technical, or material support: FC; Obtaining funding: FC, FF Supervision: FC, FF

